# Targeting the prefrontal-supplementary motor network in obsessive-compulsive disorder with intensified electrical stimulation in two dosages: A randomized, controlled trial

**DOI:** 10.1101/2023.12.01.23299285

**Authors:** Jaber Alizadehgoradel, Behnam Molaei, Khandan Barzegar Jalali, Asghar Pouresmali, Kiomars Sharifi, Amir-Homayun Hallajian, Vahid Nejati, Benedikt Glinski, Carmelo M Vicario, Michael A. Nitsche, Mohammad Ali Salehinejad

**Author notes:** corresponding authors: Mohammad Ali Salehinejad, PhD; Behnam Molaei, PhD Address: Department of Psychology and Neurosciences, Leibniz Research Centre for Working Environment and Human Factors, Ardeystr. 67, 44139 Dortmund, Germany. ***Abbreviations***: OCD = Obsessive-Compulsive Disorder; tDCS = transcranial direct current stimulation; DLPFC = dorsolateral prefrontal cortex; SMA = supplementary motor area.

## Abstract

Obsessive-Compulsive Disorder (OCD) is associated with a high disease burden, and treatment options are limited. We used intensified electrical stimulation in two dosages to target a main circuitry associated with the pathophysiology of OCD, left dorsolateral prefrontal cortex (l- DLPFC) and supplementary motor area (SMA) and assessed clinical outcomes, neuropsychological performance and brain physiology. In a double-blind, randomized-controlled trial, thirty-nine patients with OCD were randomized to sham, 2-mA or 1-mA transcranial direct current stimulation (tDCS) targeting the l-DLPFC (F3) and SMA (FC2) with anodal and cathodal stimulation respectively. The treatment included 10 sessions of 20-minute stimulation delivered twice per day with 20-min between-session intervals. Outcome measures were reduction in OCD symptoms, anxiety and depressive states, performance on a neuropsychological test battery of response inhibition, working memory, attention, and oscillatory brain activities and functional connectivity. All outcome measures except EEG were examined at pre-intervention, post-intervention and 1-month follow-up times. The 2-mA protocol significantly reduced OCD symptoms, anxiety, depression states, and improved quality of life after the intervention up to 1- month follow-up compared to the sham group, while the 1-mA protocol reduced OCD symptoms only in the follow-up and depressive state immediately after and 1-month following the intervention. Both protocols partially improved response inhibition and the 2-mA protocol reduced attention bias to OCD-related stimuli and improved reaction time in working memory performance. Both protocols increased alpha oscillatory power and the 2-mA intensified protocol decreased delta power as well. Both protocols increased connectivity in higher frequency bands at frontal-central areas compared to the sham. Modulation of the prefrontal-supplementary motor network with intensified tDCS ameliorates OCD clinical symptoms and results in beneficial cognitive effects. The 2-mA intensified stimulation resulted in larger symptom reduction and improved more converging outcome variables related to therapeutic efficacy. These results support applying the intensified prefrontal-SMA tDCS in larger trials.

## 1 Introduction

With a lifetime prevalence of 2-3%, Obsessive Compulsive Disorder (OCD) is one of the most disabling psychiatric disorders (American Psychiatric Association, 2013) with substantial functional impairment and increased risk of early mortality (Meier et al., 2016; Wayne K. Goodman et al., 2021). Individuals with OCD have unwanted and distressing thoughts (obsessions) and repetitive behaviors that the individual feels driven to perform (compulsions) (Stein et al., 2019). While cognitive-behavioral therapy with exposure/response prevention and serotonin reuptake inhibitor medication are considered first-line treatments for OCD, up to 40 % of patients fail to respond to these treatments (Romanelli et al., 2014).

Noninvasive brain stimulation techniques provide unique opportunities to not only study brain functions but also to modify core physiological parameters of human behavior and cognition (e.g., neuroplasticity) in both healthy and clinical populations (Fregni and Pascual-Leone, 2007; Polanía et al., 2018). Some noninvasive brain stimulation techniques such as repetitive transcranial magnetic stimulation (rTMS) are Food and Drug Administration (FDA)- approved for the treatment of several major neuropsychiatric disorders including OCD (Carmi et al., 2019) suggesting that other forms of techniques may be considered as a potential intervention for patients with OCD. Transcranial direct current stimulation (tDCS) is a non-invasive brain stimulation technique that uses a weak direct electrical current to modulate brain activity and excitability (Nitsche and Paulus, 2000). The exact mechanisms by which tDCS works are not fully understood, but its primary mechanism of action, which emerges immediately during stimulation involves subthreshold de- or hyperpolarization of neuronal membrane potentials, resulting in excitability-enhancing effects by anodal, and -reducing effects by cathodal stimulation in conventional protocols (Stagg and Nitsche, 2011; Polania et al., 2021). In neuropsychiatric disorders that are characterized by functional brain abnormalities (i.e., hyper- or hypoactivity of specific brain region/s and network/s), it is possible to modify altered brain functions with tDCS, and affect target behavior or cognition (Alizadehgoradel et al., 2020; Fregni et al., 2020; Salehinejad et al., 2022a; Nikolin et al., 2023). In OCD, results of tDCS studies have been mixed so far, and knowledge is still limited about optimal stimulation parameters and efficacy of interventions, such as in other clinical non-invasive brain stimulation scenarios (da Silva et al., 2019; Fregni et al., 2020; Rostami et al., 2020; Silva et al., 2021; Pinto et al., 2022).

Functional abnormalities of the dorsolateral prefrontal cortex (DLPFC) are well-documented in OCD (Li et al., 2020). Specifically, response inhibition, a core cognitive ability that is severely impaired in OCD is linked to several regions of the prefrontal cortex, including the DLPFC and inferior frontal gyrus (Aron et al., 2004, 2014; Hung et al., 2018). Another cortical region that is consistently shown to be relevant for the pathophysiology of OCD is the pre-Supplementary Motor Area (pre-SMA), which is involved in inhibitory control, especially of ongoing actions (Johansen-Berg et al., 2004; Sharp et al., 2010; Hampshire and Sharp, 2015). In OCD patients, the pre-SMA is hyperactive, especially during cognitive task performance that requires attentional and inhibitory control (Stella J. de Wit et al., 2012; Norman et al., 2019) and is, therefore, a major target of noninvasive brain stimulation treatment (Rehn et al., 2018; Gowda et al., 2019; Silva et al., 2021). Although the left DLPFC and pre-SMA have been targeted in previous tDCS studies, targeting both regions with anodal and cathodal stimulation respectively has not been reported so far (Acevedo et al., 2021; Pinto et al., 2022). Applying a protocol that can modulate the prefrontal-SMA network and presumably restore physiological abnormalities can have therapeutic effects.

Beyond the choice of the target region, stimulation parameters (e.g. stimulation intensity and repetition) are critical for the efficacy of the neurostimulation intervention, and recent work stress on optimizing and/or individualizing the intervention (Jog et al., 2019; Salehinejad et al., 2020). Physiological findings in healthy humans have shown that repeated stimulation with a short interval (e.g. two consecutive stimulation sessions with a 20 min interval) can induce long-lasting LTP-like plasticity in the brain (Monte-Silva et al., 2013). This has implications for the clinical application of tDCS. We recently showed that such a stimulation protocol, which we refer to as “intensified” protocol, has stronger and longer therapeutic effects on social anxiety disorder (Jafari et al., 2021). In the present study, we adopted the same stimulation protocol and furthermore, included different outcome variables to evaluate treatment efficacy. In addition to primary clinical symptoms, we assessed core cognitive deficits in OCD patients (e.g. response inhibition, working memory) (Robbins et al., 2019) and monitored changes in the oscillatory power spectrum and functional connectivity which are abnormally changed in OCD such as reduced and raised alpha and delta power respectively and reduced functional connectivity (Velikova et al., 2010; Wong et al., 2015; Buot et al., 2021; Perera et al., 2023).

Accordingly, in this registered, randomized, double-blind, sham-controlled clinical trial we aimed to (1) investigate the effect of intensified stimulation over prefrontal and pre-SMA regions on primary and secondary clinical variables in patients with OCD, (2) explore the stimulation dosage-dependency (1-mA vs 2-mA) of treatment efficacy, (3) and examine the effects of these interventions on cognitive (response inhibition, attention bias) and electrophysiological (oscillatory power, functional connectivity) correlates of the psychopathology of OCD. This is the first tDCS RCT in OCD that explores the effects of a novel intensified tDCS intervention at two different stimulation intensities on symptom reduction and cognitive/neural correlates of OCD.

## 2. Methods

### 2.1. Participants

This study had a randomized, double-blind, parallel-group design to prevent blinding failure and carry-over effects. Thirty-nine individuals diagnosed with OCD (mean age=31.59, SD=8.24, 26 females) were recruited from several neuropsychiatric clinics in Ardabil, Iran from August 2020 to January 2022. Patients were randomly assigned to the active and sham stimulation groups by the block randomization method (supplementary content). Sample size was calculated a priori based on a medium effect size suggested for tDCS studies (Minarik et al., 2016) (f=0.30, α=0.05, power=0.95, *N*=39, mixed-model ANOVA with 3 measurements). Two patients from the 1-mA and sham groups did not complete the whole treatment, and final analysis was conducted on 37 participants (1mA tDCS *N*=12, 2mA tDCS *N*=13, sham tDCS *N*=12) (Fig. 1). The inclusion criteria were: (1) diagnosis of OCD according to DSM-5, (2) 18-50 years old, (3) non-smoker, (4) no previous history of neurological diseases, brain surgery, epilepsy, seizures, brain damage, head injury, or metal brain implants, and (5) absence of other psychiatric disorders. Those patients taking anxiolytic (*N=6*) and/or SSRI (*N*=22) medication were receiving stable doses for 6 weeks before the experiment up to the follow-up. All participants were native speakers and had normal or corrected-to-normal vision. This was a registered clinical trial (ClinicalTrials.gov Identifier: NCT05501132) approved by the Ethics Committee of the Ardabil University of Medical Science (Ethics code: IR.ARUMS.REC.1399.102). Participants gave their written informed consent before participation (see Table 1 for demographics).

**Fig. 1:**
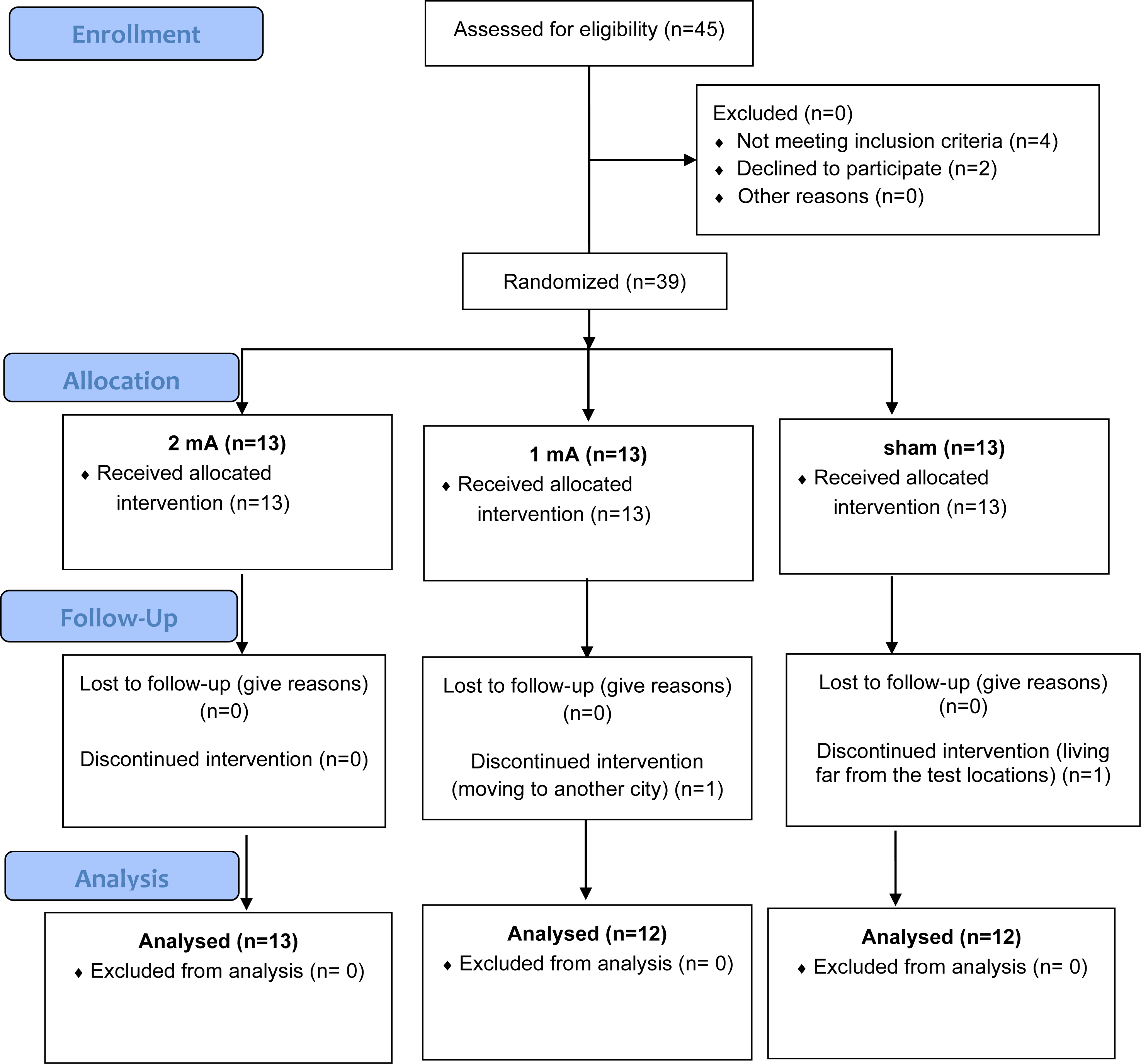
CONSORT Flow Diagram of study inclusion.

**Table 1.**
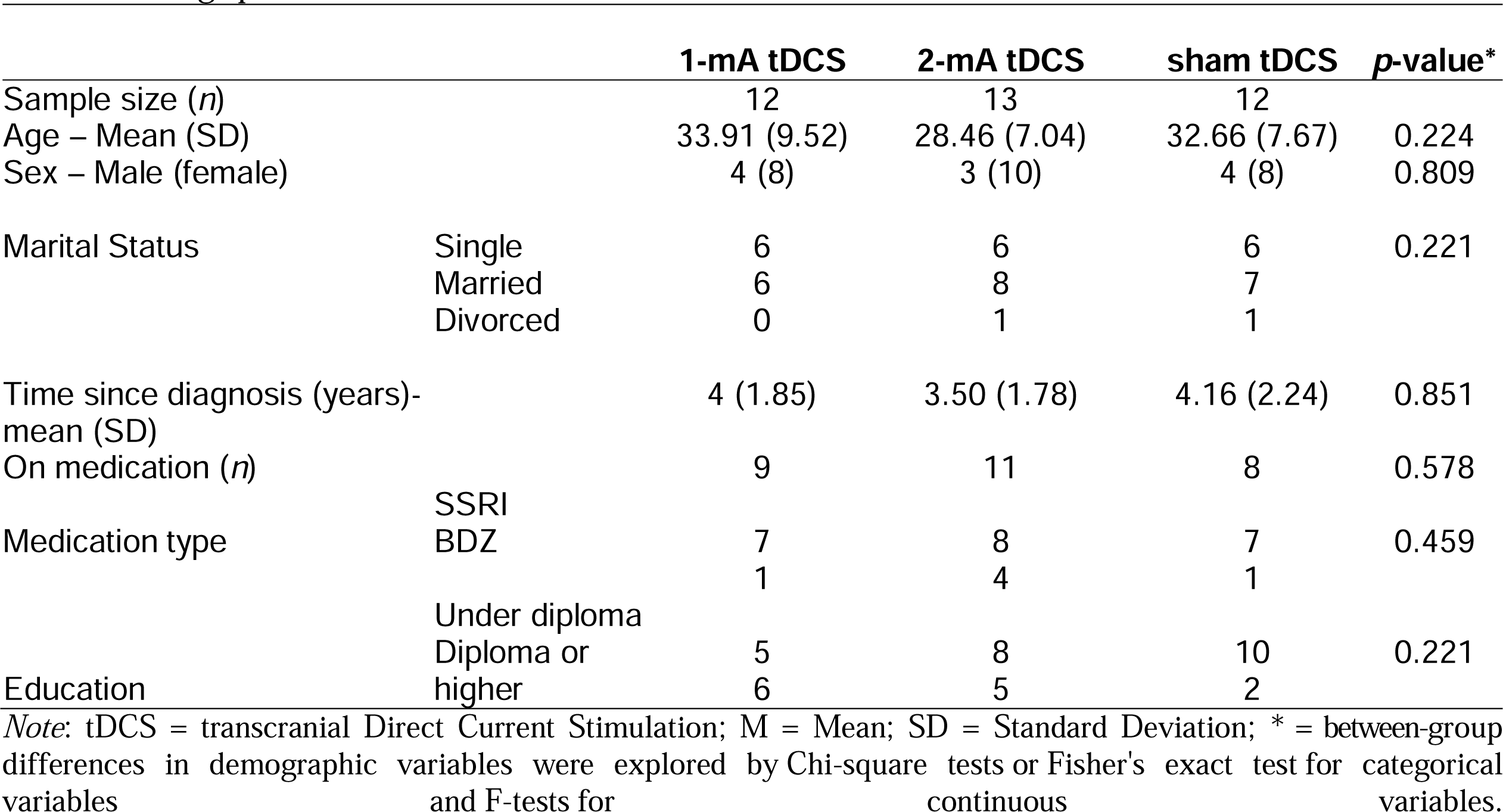
Demographic data.

|  |  | 1-mA tDCS | 2-mA tDCS | sham tDCS | p-value* |
| --- | --- | --- | --- | --- | --- |
| Sample size (*n*) |  | 12 | 13 | 12 |  |
| Age – Mean (SD) |  | 33.91 (9.52) | 28.46 (7.04) | 32.66 (7.67) | 0.224 |
| Sex – Male (female) |  | 4 (8) | 3 (10) | 4 (8) | 0.809 |
| Marital Status | Single | 6 | 6 | 6 | 0.221 |
|  | Married | 6 | 8 | 7 |  |
|  | Divorced | 0 | 1 | 1 |  |
| Time since diagnosis (years)-<br>mean (SD) |  | 4 (1.85) | 3.50 (1.78) | 4.16 (2.24) | 0.851 |
| On medication (*n*) |  | 9 | 11 | 8 | 0.578 |
| Medication type | SSRI |  |  |  | 0.459 |
|  | BDZ | 7 | 8 | 7 |  |
|  |  | 1 | 4 | 1 |  |
| Education | Under diploma |  |  |  | 0.221 |
|  | Diploma or<br>higher | 5 | 8 | 10 |  |
|  |  | 6 | 5 | 2 |  |

*Note:* tDCS = transcranial Direct Current Stimulation; M = Mean; SD = Standard Deviation; \* = between-group differences in demographic variables were explored by Chi-square tests or Fisher's exact test for categorical variables and F-tests for continuous variables.

### 2.2. Outcome measures (primary and secondary clinical measures, cognitive deficits and brain physiology)

#### 2.2.1. primary and secondary clinical measures

The primary outcome measure to examine the effects of the intervention on OCD symptoms was the Yale-Brown Obsessive-Compulsive Scale (Y-BOCS) (Goodman et al., 1989). Additionally, anxiety and depressive states were tested by the Beck Anxiety Inventory (BAI) (Steer and Beck, 1997) and the Beck Depression Inventory (BDI-II) (Beck et al., 1961), respectively, and quality of life was assessed with the WHO Quality of Life Questionnaire (WHOQUL) (Skevington et al., 2004). These measures were used to evaluate the clinical efficacy of the intervention. The Y-BOCS is the most widely used clinician-rated interview for assessing OCD symptom severity and is a reliable measure of treatment-based reduction of symptoms (Maust et al., 2012). The BAI is also well suited to monitor treatment outcomes (Leyfer et al., 2006), and the evaluated anxiety state is correlated with OCD symptoms (Velloso et al., 2016; Reuman et al., 2017). Similarly, BDI-II scores are associated with OCD symptoms (Velloso et al., 2016), in line with the fact that around one-third of OCD patients suffer from comorbid depression (Overbeek et al., 2002). A detailed description of these measures can be found in the supplementary information.

#### 2.2.2. Cognitive assessment and brain physiology

We used a battery of neuropsychological tests that are sensitive to the cognitive deficit profile of OCD affected by interventions. Deficits of inhibitory control, working memory performance, and attention (e.g., sustained attention, set-shifting) are among the most well-documented cognitive deficits in OCD (Shin et al., 2013; Benzina et al., 2016; Robbins et al., 2019). Importantly, these cognitive deficits are associated with frontal–striatal and frontal dysfunctions (van den Heuvel et al., 2005; Norman et al., 2016; Heinzel et al., 2018; Norman et al., 2019) which were targeted by the intervention in this experiment. We examined response inhibition with the Go/No-Go task and Flanker test, working memory with the n-back task, and attention bias to COD-related stimuli with an adapted dot probe task. A detailed description of these measures is provided in the supplementary information. Finally, we monitored resting-EEG to see how power spectrum and functional connectivity are changed after the intervention specifically in frequency bands of interest (e.g. alpha, delta, gamma) (Velikova et al., 2010; Wong et al., 2015; Buot et al., 2021; Perera et al., 2023). A detailed description of the measures and EEG data preprocessing and analysis are in the supplementary content.

### 2.3. tDCS

Direct currents were generated by an electrical stimulator (Oasis Pro, Mind Alive, Canada), and applied through a pair of saline-soaked sponge electrodes (7×5 cm) for two periods of 20 min and 20 min intervals between each stimulation period (Jafari et al., 2021). Stimulation was delivered on 5 consecutive days (two stimulations per day). In both active (1-mA, 2-mA) and sham conditions, anodal and cathodal electrodes were placed over the left DLPFC (F3), and right pre-SMA (FC2) respectively, to guarantee a minimum 6 cm distance between the edges of the electrodes (Woods et al., 2016). To localize the right pre-SMA first, the pre-SMA was identified using the EEG 10–20 system for electrode positioning (i.e., 15% cm anterior to Cz) (D’Urso et al., 2016; Silva et al., 2021). In sham stimulation, the electrical current was ramped up and down for 30 seconds to generate the same sensation as in the active condition and then turned off (Brunoni et al., 2014). To guarantee blinding, tDCS was applied by independent investigators who were not involved in outcome measures rating (Gandiga et al., 2006). A side-effect survey was done after each tDCS session. Blinding efficacy was not explored among patients and experimenters. A 3D model of the current flow in the head was created to determine induced electrical fields in the brain for the above-mentioned tDCS protocol (Fig. 2).

**Fig. 2:**
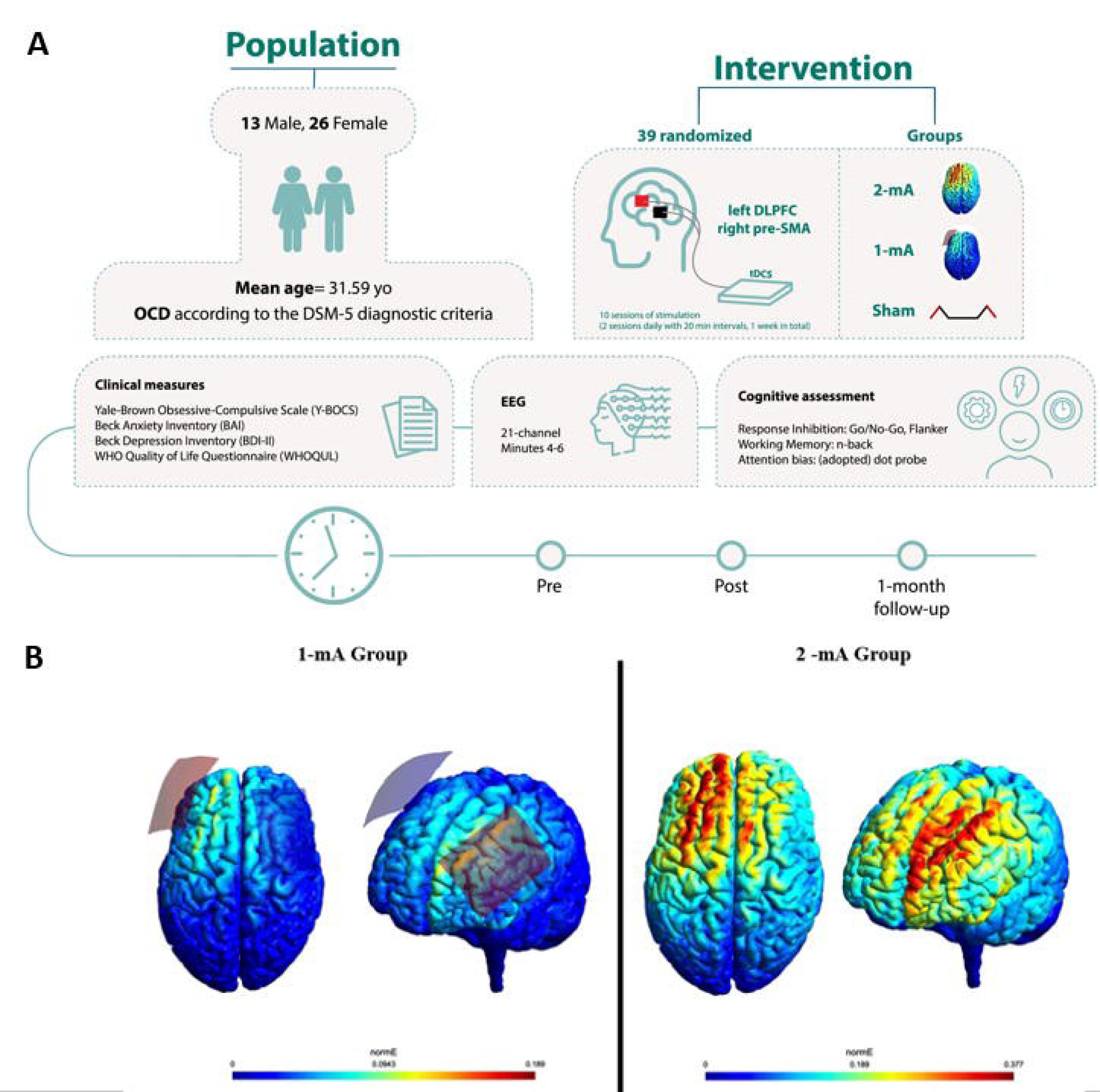
**A**, Experimental procedure. The experiment was conducted in a randomized, double-blind, sham-controlled parallel-group design. Participants were randomized to 3 tDCS arms: 1- mA tDCS (n= 12), 2-mA tDCS (n= 13), and sham tDCS (n= 12). **B**, Results of the electrical field simulation for the current flow in the head based on the applied protocols. The anodal electrode (red) was placed over the left DLPFC and the cathode (blue) over the pre-SMA (FC2). The induced electric fields (EFs) were calculated using ROAST (Huang et al., 2019), an open-source pipeline for transcranial electrical stimulation (tES) modeling (available at www.parralab.org/roast/). ROAST employs SPM12, iso2mesh, and getDP for segmentation of MRI, finite element meshing, and solving the finite element model to estimate the EF distribution in the head. Simulations were performed for the MNI-152 standard head (Grabner et al., 2006). Two stimulation protocols were modeled: one with 7×5 cm electrodes over the left DLPFC (F3) and the pre-SMA (FC2), with 1-mA current intensity, and the same parameters except for the intensity which was 2-mA (10-05 EEG electrode positioning system). The average EF value (undirected) in each region was calculated based on the sum of EF values and the number of voxels in the area. All the calculations were performed using FSL and Matlab.

### 2.4. Procedure

Prior to the experiment, participants completed a brief questionnaire to evaluate their suitability for brain stimulation. All participants received 10 sessions of stimulation (2 sessions daily, 5 days in total) with 20-minute intervals between the sessions. To avoid confounding effects of the intervention at circadian non-preferred time, which can significantly affect neuroplasticity induction (Salehinejad et al., 2021b), all stimulation sessions took place between 11:00-14:00 and participants were not under sleep pressure (Salehinejad et al., 2022b). Clinical cognitive measures were evaluated before the first intervention (pre-intervention), right after the end of the last intervention (post-intervention), and 1-month following the last stimulation session (follow-up). EEG measurements took place only before and after the intervention.

Patients were instructed about the tasks before the beginning of the experiment. None of the patients received any kind of psychotherapy during the study. Participants were blind to the study hypotheses and stimulation conditions. The experimenter who conducted the outcome measures was blinded to the tDCS conditions (Fig. 2).

### 2.5. Statistical analysis

Data analyses were conducted with the statistical package SPSS, version 26.0 (IBM, SPSS, Inc., Chicago, IL) and the GraphPad Prism 8.2.1 (GraphPad Software, San Diego, California). The normality and homogeneity of data distribution, and variance were confirmed by Shapiro-Wilk and Levin tests, respectively. Between-group differences in demographic variables were explored by Chi-square tests or Fisher’s exact test for categorical variables and F-tests for continuous variables. A multivariate Analysis of Variance (MANOVA) was first performed on the post-intervention and follow-up means of all outcome variables with group as the fixed factor and pre-intervention measures as covariates. This was to help protect against inflating the Type 1 error rate in the follow-up ANOVAs and post-hoc comparisons. A series of one-way ANOVA’s on each dependent variable was conducted as follow-up tests to the MANOVA. Finally, a series of post-hoc analyses were calculated using the Tukey’s multiple comparisons tests to examine individual mean difference comparisons across groups (active 1mA, active 2mA, sham) and time points (pre-intervention, post-intervention, follow-up). The critical level of significance was 0.05 for all statistical analyses.

## 3. Results

### 3.1. Side effects and baseline assessment

Participants tolerated the stimulation well and no adverse effects were reported during and after stimulation. No significant difference was found between the group ratings of tDCS side effects (Supplementary Table S1). No significant between-group differences emerged in the pre-intervention measurements (Supplementary Table S2).

#### 3.2. Primary clinical outcome: Reduction of OCD symptoms and anxiety

A statistically significant MANOVA effect was obtained for both post-intervention (Pillais’ Trace = 1.64, *F*_(24, 24)_ = 4.63, *p*<0.001) and follow-up measurements (Pillais’ Trace = 2.98, *F*_(24, 24)_ = 4.63, *p*=0.005). The results of the follow-up ANOVAs revealed a significant main effect of group on both Y-BOCS scores (post-intervention: *F*_(2, 22)_=7.14, *p*=0.004, η*p^2^*=0.394; follow-up: *F*_(2, 22)_=13.54, *p*<0.004, η*p^2^*=0.552) and BAI scores (post-intervention*: F*_(2, 22)_ = 8.78, *p*=0.002, η*p^2^*=0.423; follow-up: *F*_(2, 22)_=5.78, *p*=0.010, η*p^2^*=0.345). Next, Tukey’s multiple test comparisons were performed on individual mean difference and showed a significant decrease in Y-BOCS scores at the post-intervention time in the 2-mA group (*p*=0.021, *d=*0.98), at the 1- month follow-up in both the 2-mA (*p*=0.004, *d=*1.01) and 1-mA (*p*=0.013, *d=*1.23) groups, but no significant changes in the sham group as compared to pre-intervention time (Fig. 3A). When compared to the sham group, reduced Y-BOCS scores were significant in both active groups only at the follow-up (1-mA: *p*=0.044, *d=*1.16; 2-mA: *p*=0.045, *d=*0.82) (Fig. 3B). For the BAI scores, Tukey’s multiple test comparisons showed a significant decrease in BAI scores from pre-intervention to both post-intervention (*p*=0.025, *d=*0.85) and 1-month follow-up (*p*=0.009, *d=*0.97) only in the 2-mA group (Fig 3C). When compared to the sham group, both active groups showed a non-significant trendwise reduction of BAI scores at the post-intervention assessment (1-mA: *p*=0.074; 2-mA: *p*=0.083) (Fig. 3D).

**Fig. 3:**
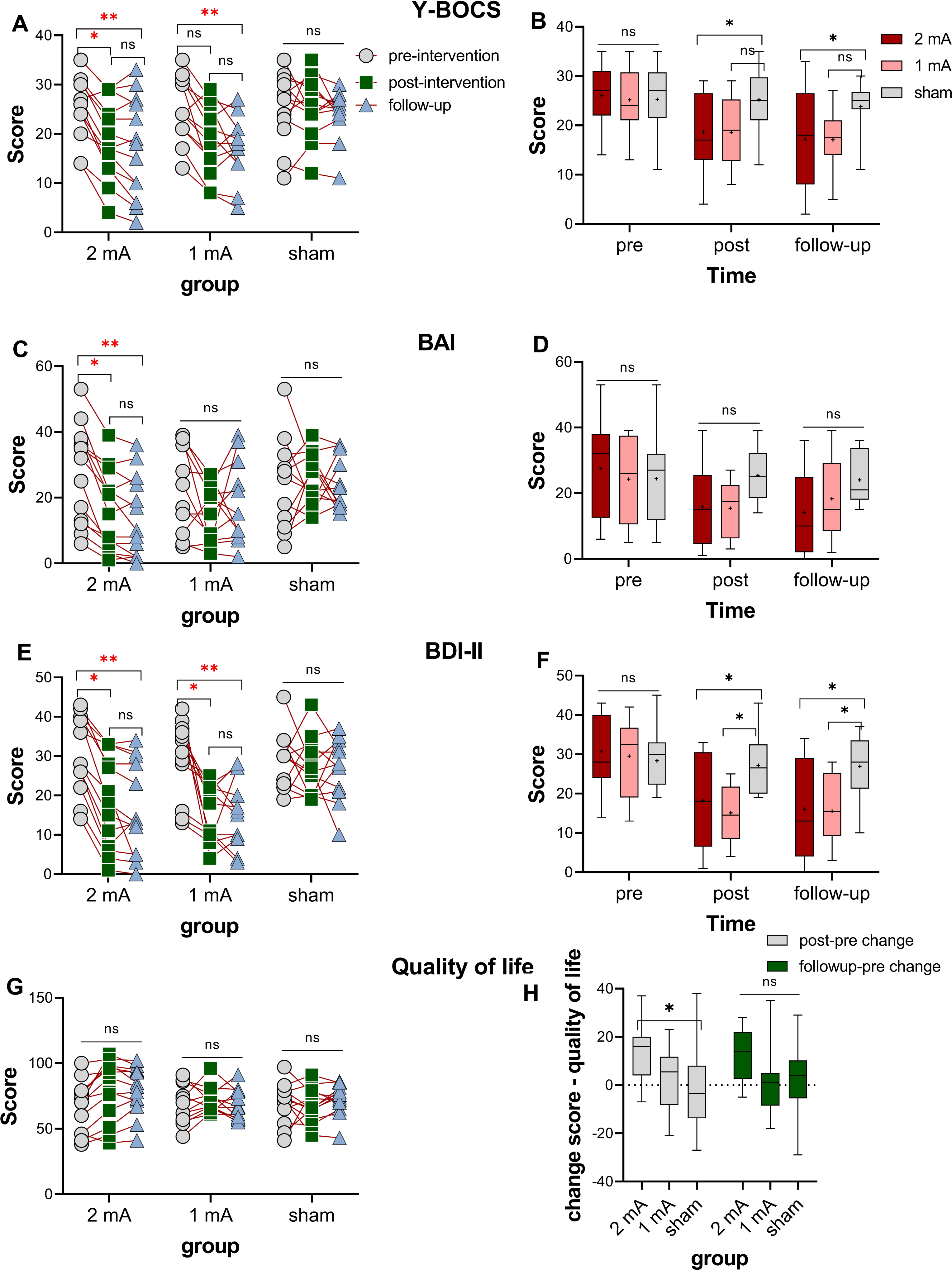
OCD symptoms measured by Y-BOCS (**A, B**) and treatment-related variables (**C-H**) (anxiety measured by BAI, depressive state measured by BDI-II, and quality of life measured by the WHO quality of life scale), before and immediately after intervention and 1-month follow-up. *Note*: Y-BOCS = Yale-Brown Obsessive-Compulsive Scale; BAI = Beck Anxiety Inventory; BDI-II = Beck Depression Inventory-II. Floating asterisks [*] in the left panel represent a significant difference between pre-intervention measurement vs post-intervention and follow-up measurements in all groups. Floating asterisks [*] in the right-side figures indicate a significant difference between active stimulation groups (1 and 2-mA) vs sham tDCS at each time point. Ns = nonsignificant. All pairwise comparisons were conducted using Tukey’s multiple-test comparisons. All error bars represent s.e.m

### 3.3. Secondary clinical outcomes: mood and quality of life

The results of the follow-up ANOVAs revealed a significant main effect of group on BDI-II scores (post-intervention: *F*_(2, 22)_=7.13, *p*=0.004, η*p^2^*=0.394; follow-up: *F*_(2, 22)_=10.47, *p*<0.001, η*p^2^*=0.488) and quality of life (post-intervention: *F*_(2, 22)_=3.58, *p*=0.045, η*p^2^*=0.246; follow-up: *F*_(2, 22)_=7.33, *p*=0.004, η*p^2^*=0.400). Tukey’s multiple test comparisons showed that BDI-II scores were reduced from the pre-intervention to both post-intervention and 1-month follow-up assessment in both, 2-mA (post-intervention: *p*=0.002, *d=*1.15; follow-up: *p*<0.001, *d=*1.34) and 1-mA (post-intervention: *p*<0.001, *d=*1.65; follow-up: *p*=0.001, *d=*1.49) groups, but not in the sham group, and reduced depressive state was significant in both active groups vs. the sham group (Fig. 3E, F). No significant individual mean differences were found across groups in quality of life scores (Fig. 3G). However, we calculated the changes in quality of life scores from the baseline to post-intervention and follow-up. Tukey’s multiple test comparisons of score changes across groups showed that quality of life scores significantly improved after the intervention only in the 2 mA group (Fig. 3H).

### 3.4. Cognitive functions: Improved inhibitory control in both active tDCS groups

In the Flanker test, the follow-up ANOVAs revealed a significant main effect of group on both, congruent (post-intervention: *F*_(2, 22)_=10.08, *p*<0.001, η*p^2^*=0.478; follow-up: *p*=0.901) and incongruent trials (post-intervention: *F*_(2, 22)_=8.01, *p*=0.002, η*p^2^*=0.422; follow-up: *p*=0.445) only after the intervention and not follow-up. Tukey’s multiple test comparisons revealed a significant *pre* vs *post*-intervention RT reduction of incongruent stimuli (*p*=0.021, *d=*1.35) only in the 1- mA group which, however, was not significant vs the sham (Fig. 4A, B). In the Go/No-Go task, No-Go trial performance, the results of the follow-up ANOVAs revealed a significant main effect of group for No-Go trials reaction time (post-intervention: *F*_(2, 22)_=7.11, *p*=0.004, η*p^2^*=0.393; follow-up: *F*_(2, 22)_=4.34, *p*=0.026, η*p^2^*=0.283) and a marginally significant effect on accuracy at the post-intervention measurement (*F*_(2, 22)_=3.39, *p_follow-up_*=0.054, η*p^2^*=0.233).

**Fig. 4:**
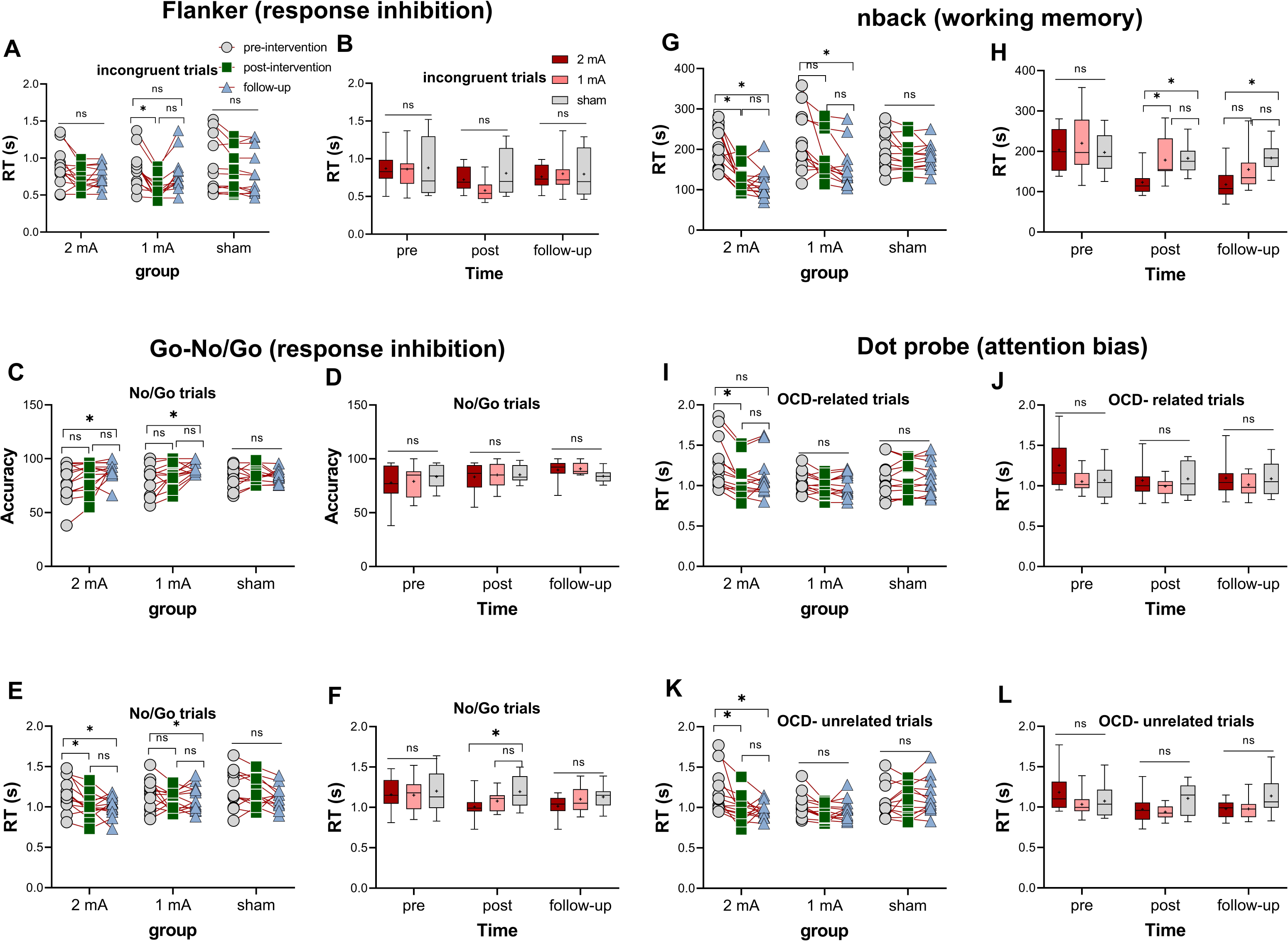
Response inhibition measured by Flanker (**A-B**) and Go-No/Go (**C-F**) tasks, before, immediately after the intervention, and at the 1-month follow-up. Working memory and attention bias were measured by n-back (**G-H**) and dot-probe (**I-L**) tasks, before, immediately after the intervention, and 1-month follow-up. *Note*: RT = Reaction time; s = seconds. Floating asterisks [*] in the left panel represent a significant difference between pre-intervention measurement vs post-intervention and follow-up measurements in all groups. Floating asterisks [*] in the right panel indicate a significant difference between active stimulation groups (1 and 2-mA) vs sham tDCS at each time point. ns = nonsignificant. All pairwise comparisons were conducted using Tukey’s multiple-test comparisons. All error bars represent s.e.m

Tukey’s multiple test comparisons showed increased accuracy from the pre-intervention to the follow-up measurement in 2-mA (*p*=0.022, *d=*0.87) and 1-mA (*p*=0.032, *d=*0.27) groups (Fig 4C). The 2-mA protocol significantly reduced RT from *pre* vs *post*-intervention (*p*=0.027, *d=*1.92) and *pre* vs *follow-up* (*p*=0.037, *d=*1.95) as well (Fig. 4E) and here the performance speed on the No-Go trials was significantly faster in the 2-mA group vs the sham after the intervention (*p*=0.014, *d=*1.73) (Fig. 4F).

### 3.5. Cognitive functions: working memory and attention bias

In working memory performance, the follow-up ANOVAs revealed a significant main effect of group only on performance speed after the intervention (*F*_(2, 22)_=16.76, *p*<0.001, η*p* =0.604) and 1-month follow-up (*F*_(2, 22)_=13.87, *p*<0.001, η*p* =0.558). Tukey’s multiple test comparisons showed a significantly faster *pre*- vs post-intervention RT (*p*<0.001, *d=*1.92) and *pre* vs *follow-up* RT (*p*<0.001, *d=*1.95) in the 2-mA and a significant *pre* vs *follow-up* RT reduction (*p*=0.003, *d=*1.01) in the 1-mA group. In the 2-mA group, this RT reduction was furthermore larger than that of the sham group at post-intervention (*p*=0.007, *d=*1.73) and follow-up (*p*=0.002, *d=*1.91) measurements (Fig. 4G, H). Finally, in the attention bias of patients to OCD-related stimuli, the follow-up ANOVAs showed a significant main effect of group for both, OCD-related (post-intervention: *F*_(2, 22)_=8.82, *p*=0.002, η*p^2^*=0.445; follow-up: *F*_(2, 22)_=5.53, *p*=0.011, η*p^2^*=0.335) and unrelated stimuli (post-intervention: *F*_(2, 22)_=5.82, *p*=0.009, η*p^2^*=0.346; follow-up: *F*_(2, 22)_=9.45, *p*=0.001, η*p^2^*=0.462). Tukey’s multiple test comparisons showed a significantly faster *pre*- vs *post*-intervention RT for both, OCD-related (*p*<0.026, *d=*0.70) and unrelated (*p*<0.010, *d=*0.93) stimuli in the 2-mA group (Fig. 4I, K). No significant between-group RT differences were however found for the post-intervention and follow-up measurements.

### 3.6. Intervention-related changes in EEG power spectrum density and functional connectivity

PSD Analysis with a cluster-based permutation test (post vs pre) revealed a significant increase in relative alpha power in the left frontal region (cluster-level statistic = 16, *p*<0.01) and the occipital region (cluster-level statistic = 11, *p*<0.05) in the 2-mA group compared to the sham group. Additionally, a significant decrease in relative delta power was observed in a cluster located in the occipital region (cluster-level statistic = -11, *p*<0.05) in the 2-mA group compared to the sham group. In the 1-mA group, a significant increase in relative alpha power was observed in the right frontal region (cluster-level statistic = 11, *p*<0.001) (Fig. 5C). Regarding functional connectivity, comparative analysis of post-intervention Phase Locking Value (PLV) matrices showed a general trend of increased connectivity in higher frequency bands in both active groups as compared to the sham group (Fig. 5A, B). When we compared both groups with each other, the 2-mA group generally decreased functional connectivity across most frequency bands (EEG connectivity results are fully described in the supplementary content). We did not see any interesting correlation between EEG parameters and clinical/cognitive measures.

**Fig. 5:**
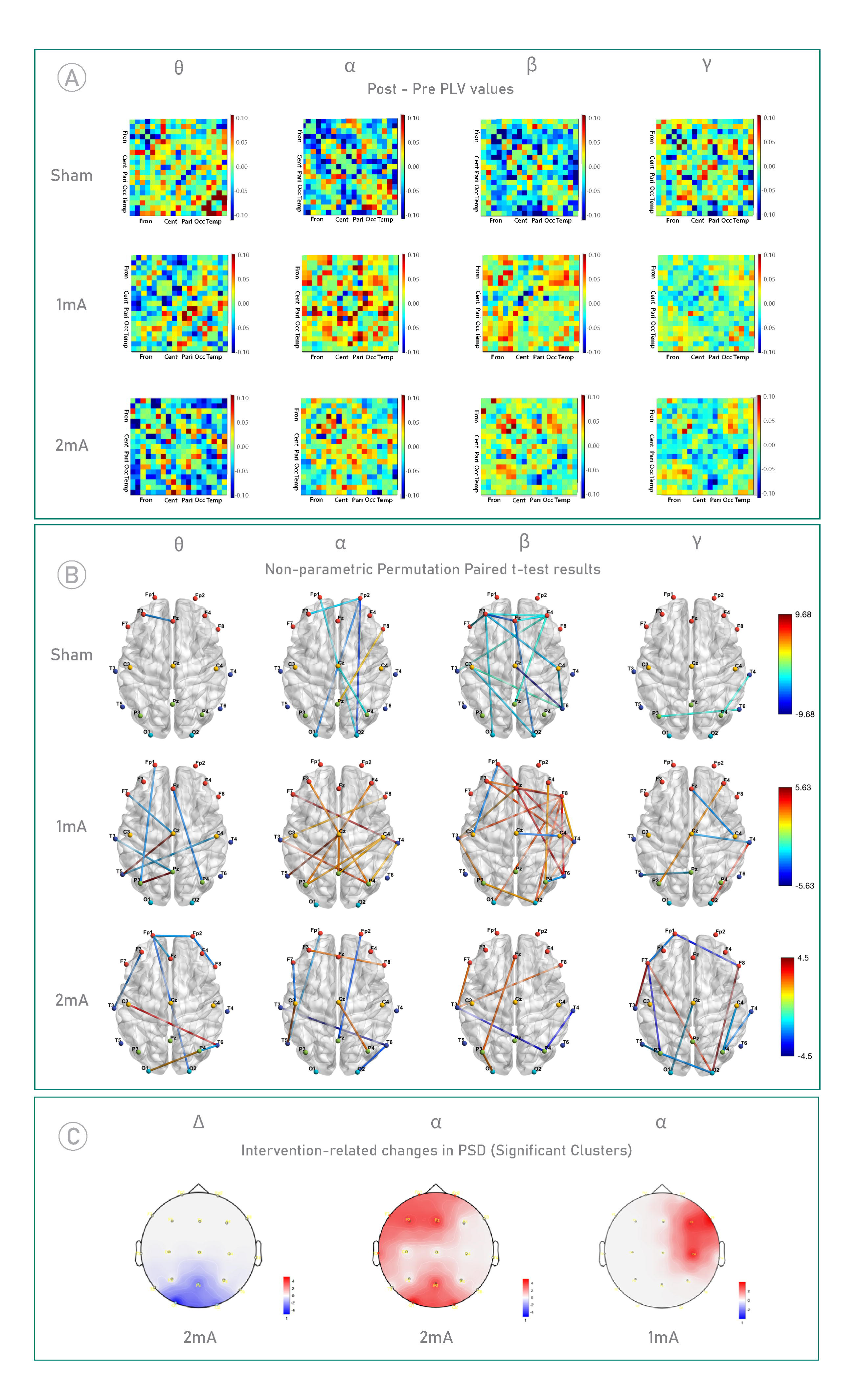
Functional connectivity and Power spectrum density (PSD) analyses: **A,** Post-minus-pre tDCS intervention connectivity matrices (PLV) averaged across participants in each intervention group for theta, alpha, beta, and gamma frequencies. Only the first four letters of the respective scalp regions are used for abbreviation: Frontal (Fron), Central (Cent), Parietal (Pari), Occipital (Occ), Temporal (Temp). **B,** Visualization of the edge-level data with significant subnetworks in each frequency band. The size and color of the edges are determined by the t-value. Red edges represent an increase in functional connectivity between respective nodes and blue edges represent a decrease in functional connectivity between respective nodes. Node colors represent specific regions- Frontal (Red), Central (Yellow), Parietal (Green), Occipital (Cyan), and Temporal (Dark Blue). Associated electrodes within each region are presented in supplementary materials. **C**, Intervention-related changes in PSD. A cluster-based permutation test was conducted on relative PSD values (post-intervention minus pre-intervention) within each active group in comparison to the sham group, identifying significant clusters. The left panel represents a statistically significant decrease in relative delta power in the 2-mA group compared to the sham group (cluster-level statistic = -11, *p*<0.05). The middle panel illustrates an increase in relative alpha power in the 2-mA group compared to the sham group in the left frontal region (cluster-level statistic = 16, *p*<0.01), and occipital region (cluster-level statistic = 11, *p*<0.05). The right panel displays a marked increase in relative alpha power in the 1-mA group relative to the sham group, with a significant cluster identified in the right frontal region (cluster-level statistic = 11, *p*<0.05)

## 4. Discussion

In this randomized, double-blind, sham-controlled, parallel-group clinical trial, we investigated the impact of an intensified tDCS protocol (stimulation twice per day with 20-min intervals) over the prefrontal-supplementary motor network, in two dosages (1-mA vs 2-mA) on primary clinical symptoms, neuropsychological performance, and electrophysiological correlates in patients with OCD. The 2-mA stimulation dosage significantly reduced OCD symptoms and anxiety after the intervention and in the follow-up. Both active stimulation protocols significantly reduced depressive symptoms. At the neuropsychological level, both active protocols partially improved response inhibition and the 2-mA protocol reduced attention bias to threat-related stimuli and improved working memory performance as well. Both protocols increased alpha and the 2-mA protocol decreased theta oscillatory power too. Both protocols increased connectivity in higher frequency bands at frontal-central areas compared to the sham. No significant changes were observed in the sham group for any outcome measures.

These findings can be explained from neurophysiological and neuropsychological perspectives. The hallmark finding of neuroimaging studies refers to a lateral hypoconnectivity (including the DLPFC) and medial hyperconnectivity (including the pre-SMA) in OCD (van den Heuvel et al., 2016; Robbins et al., 2019), which was the rationale for applying our stimulation protocol. We applied anodal stimulation over the left DLPFC to increase the activity of this region and cathodal stimulation of the pre-SMA to downregulate activity. With causal modulation of cerebral excitability with tDCS (Polanía et al., 2018), we expected to restore functional abnormalities in the OCD-relevant brain circuitry, and in principal accordance, this intervention was associated with behavioral and clinical improvement in this study. In further accordance, the intervention especially after 2-mA stimulation restored altered alpha and delta oscillatory power in patients (Perera et al., 2023) and both protocols increased connectivity in the prefrontal regions, which is reduced in OCD patients, and thus likely an appropriate treatment target (Li et al., 2020).

In addition to neurophysiological changes, neuropsychological accounts could also explain our findings. The most well-known psychological mechanisms underlying OCD psychopathology include impaired cognitive control (the inability to regulate compulsive behavior) (Menzies et al., 2007), impaired cognitive flexibility (the inability to regulate thinking) (Chamberlain et al., 2007), and impaired balance between goal-directed behavior and more automatic habit learning (Graybiel and Rauch, 2000; Everitt and Robbins, 2016). Importantly, these cognitive abilities are related to lateral and medial prefrontal cortices (Aron et al., 2014; Robbins et al., 2019; Salehinejad et al., 2021a). The behavioral tasks we used are primarily related to cognitive control and cognitive flexibility (e.g. response inhibition, working memory), and performance of these tasks was significantly improved after intervention, more obviously in the 2-mA group. Here, it should however be noted that the effects of both protocols on response inhibition were smaller than expected, which could be due to the higher relevance of the right prefrontal region in cognitive inhibition (Schroeder et al., 2020). That said, anodal stimulation of the left DLPFC was shown to improve executive functions in neuropsychiatric patients also in previous studies (Alizadehgoradel et al., 2020; Jafari et al., 2021; Basharpoor et al., 2022), and might explain treatment effects in OCD patients.

One major rationale of this study was to identify the effect of different stimulation dosages on treatment efficacy, specifically in the intensified protocol which we already applied in another study with promising results (Jafari et al., 2021). This protocol has not been explored before in OCD to the best of our knowledge. Our results in this study show that the 2-mA intensified tDCS protocol was overall more effective than both, sham stimulation and the 1-mA protocol, especially for the clinical variables, and it improved more outcome measures including measures of behavioral performance compared to the 1-mA protocol (e.g., working memory, attention bias). The rationale behind the protocol comes from a study showing that twice stimulation with 20 min intervals leads to longer aftereffects on cortical excitability compared to non-repeated stimulation or stimulation with long intervals, and resembles features of late-phase LTP (Monte-Silva et al., 2013; Agboada et al., 2020). This finding has at least two important clinical implications. First, the 2-mA stimulation is associated with higher clinical efficacy in OCD, and probably in other anxiety disorders, as shown in our previous work in patients with social anxiety disorder (Jafari et al., 2021). Second, the intensified stimulation (twice per day with a 20 min interval), has significant clinical efficacy for OCD symptoms and treatment-related variables. This is in line with physiological studies that have shown that repeated tDCS sessions induce larger increases in excitability (Ho et al., 2016) and more importantly suggest that the intensified protocol (repetition of two 20-minute stimulation with 20-minute interval between) can be promising for clinical application in other neuropsychiatric diosders.

Our protocol was different from other commonly applied protocols in other aspects. First and to the best of our knowledge, none of the previous randomized trials targeted the prefrontal- SMA network by stimulating both left DLPFC and pre-SMA (Gao et al., 2022; Camacho-Conde et al., 2023). Additionally, this is also the first randomized-controlled trial that compared the efficacy of two stimulation dosages which is typically needed for establishing clinical efficacy of an intervention. Finally, in comparison to other protocols used in previous studies, a recent metanalysis of tDCS RCTs in OCD showed that protocols that applied cathodal stimulation over the pre-SMA with an extracephalic return electrode delivered stronger electric fields to the circuity involved in OCD in comparison to the other montages (Pinto et al., 2022). None of these tDCS studies targeted the left DLPFC with anodal tDCS. This metanalysis however did not find significant differences between active vs sham tDCS in contrast to our study.

Our study had several limitations. First, the intrinsically limited focality of tDCS can result in a relatively diffuse stimulation. Neuroimaging methods will help to more accurately identify the regions directly affected by tDCS in future studies. Furthermore, we did not examine blinding efficacy in patients and could not measure EEG in the follow-up due to COVID-19- related restrictions. With respect to blinding efficacy, the 2 mA intensity typically results in more sensations over the skin as compared to the sham and 1 mA protocol which may affect patients’ blinding. However, there was no significant difference in reported ratings of tDCS side effects between groups (see supplementary information, Table S1).

Taken together, our findings suggest that the intensified prefrontal-supplementary motor cortex tDCS protocol introduced for the treatment of OCD is promising and might be effective in other neuropsychiatric disorders. Both, primary OCD symptoms and secondary treatment-related variables (anxiety, depressive state) improved after the intervention, especially in the 2-mA group. Response inhibition was partially improved by both intensified protocols, but working memory, and attentional bias to OCD-related stimuli improved after the 2-mA intervention only. Partial effects of the intervention on response inhibition might suggest further optimizing the protocol by targeting the right prefrontal cortex which was not the primary target here and the sessions were relatively low. Both protocols significantly restored brain oscillatory power in the frequency bands introduced as biomarkers of OCD. Future larger trials with longer follow-up assessments are needed to support clinical efficacy of this intervention.

## Data Availability

All data produced in the present study are available upon reasonable request to the authors

## Acknowledgments

MAN is supported by the Deutsche Forschungsgemeinschaft (DFG, German Research Foundation)—Project Number 316803389—SFB 1280, project A6, and by the German Centre of Mental Health (Project Number 01EE2302D).

## Conflict of Interest Disclosures

MAN is a member of the Scientific Advisory Boards of Neuroelectrics and NeuroDevice. All other authors declare no competing interests.

## Author Contributions

**Jaber Alizadehgoradel**: Conceptualization, Investigation, Data curation, Validation. **Behnam Molaei:** Resources, Project administration, Data curation. **Khandan Barzegar Jalali** & **Asghar Pouresmali**: Investigation, Data curation. **Amir-Homayoun Hallajian & Kiomars Sharifi**: EEG analysis, 3D Modeling, visualization, Writing – original draft (EEG part). **Vahid Nejati**: Software. **Benedikt Glinski**: Data analysis. **Carmelo M. Vicario:** Writing - review & editing. **Michael A. Nitsche**: Supervision, Methodology, Writing - review & editing. **Mohammad Ali Salehinejad**: Conceptualization, Methodology, Supervision, Writing – original draft, Writing - review & editing, Formal analysis.

## Notes

### Clinical Trial

NCT05501132

### Author Declarations

This was a registered clinical trial (ClinicalTrials.gov Identifier: NCT05501132) approved by the Ethics Committee of the Ardabil University of Medical Science (Ethics Code: IR.ARUMS.REC.1399.102).

## References

1. Acevedo N, Bosanac P, Pikoos T, Rossell S, Castle D (2021) Therapeutic Neurostimulation in Obsessive-Compulsive and Related Disorders: A Systematic Review. Brain Sciences 11:948.

2. Agboada D, Mosayebi-Samani M, Kuo M-F, Nitsche MA (2020) Induction of long-term potentiation-like plasticity in the primary motor cortex with repeated anodal transcranial direct current stimulation – Better effects with intensified protocols? Brain Stimulation 13:987–997.

3. Alizadehgoradel J, Nejati V, Sadeghi Movahed F, Imani S, Taherifard M, Mosayebi-Samani M, Vicario CM, Nitsche MA, Salehinejad MA (2020) Repeated stimulation of the dorsolateral-prefrontal cortex improves executive dysfunctions and craving in drug addiction: A randomized, double-blind, parallel-group study. Brain Stimulation 13:582–593.

4. American Psychiatric Association (2013) Diagnostic and statistical manual of mental disorders (DSM-5®): American Psychiatric Pub.

5. Aron AR, Robbins TW, Poldrack RA (2004) Inhibition and the right inferior frontal cortex. Trends in Cognitive Sciences 8:170–177.

6. Aron AR, Robbins TW, Poldrack RA (2014) Inhibition and the right inferior frontal cortex: one decade on. Trends in Cognitive Sciences 18:177–185.

7. Basharpoor S, Zakibakhsh Mohammadi N, Heidari F, Azarkolah A, Vicario CM, Salehinejad MA (2022) Emotional working memory training improves cognitive inhibitory abilities in individuals with borderline personality trait: A randomized parallel-group trial. Journal of Affective Disorders 319:181–188.

8. Beck AT, Ward CH, Mendelson MM, Mock JJ, Erbaugh JJ (1961) An inventory for measuring depression. Archives of General Psychiatry 4:561–571.

9. Benzina N, Mallet L, Burguière E, N’Diaye K, Pelissolo A (2016) Cognitive Dysfunction in Obsessive-Compulsive Disorder. Current Psychiatry Reports 18:80.

10. Brunoni AR, Schestatsky P, Lotufo PA, Benseñor IM, Fregni F (2014) Comparison of blinding effectiveness between sham tDCS and placebo sertraline in a 6-week major depression randomized clinical trial. Clin Neurophysiol 125:298–305.

11. Buot A, Karachi C, Lau B, Belaid H, Fernandez-Vidal S, Welter M-L, Mallet L (2021) Emotions Modulate Subthalamic Nucleus Activity: New Evidence in Obsessive-Compulsive Disorder and Parkinson’s Disease Patients. Biological Psychiatry: Cognitive Neuroscience and Neuroimaging 6:556–567.

12. Camacho-Conde JA, del Rosario Gonzalez-Bermudez M, Carretero-Rey M, Khan ZU (2023) Therapeutic potential of brain stimulation techniques in the treatment of mental, psychiatric, and cognitive disorders. CNS Neurosci Ther 29:8-23.

13. Carmi L, Tendler A, Bystritsky A, Hollander E, Blumberger DM, Daskalakis J, Ward H, Lapidus K, Goodman W, Casuto L, Feifel D, Barnea-Ygael N, Roth Y, Zangen A, Zohar J (2019) Efficacy and Safety of Deep Transcranial Magnetic Stimulation for Obsessive-Compulsive Disorder: A Prospective Multicenter Randomized Double-Blind Placebo-Controlled Trial. Am J Psychiatry 176:931–938.

14. Chamberlain SR, Fineberg NA, Menzies LA, Blackwell AD, Bullmore ET, Robbins TW, Sahakian BJ (2007) Impaired Cognitive Flexibility and Motor Inhibition in Unaffected First-Degree Relatives of Patients With Obsessive-Compulsive Disorder. American Journal of Psychiatry 164:335–338.

15. D’Urso G, Brunoni AR, Mazzaferro MP, Anastasia A, de Bartolomeis A, Mantovani A (2016) Transcranial direct current stimulation for obsessive–compulsive disorder: a randomized, controlled, partial crossover trial. Depression and anxiety 33:1132–1140.

16. da Silva RdMF, Batistuzzo MC, Shavitt RG, Miguel EC, Stern E, Mezger E, Padberg F, D’Urso G, Brunoni AR (2019) Transcranial direct current stimulation in obsessive-compulsive disorder: an update in electric field modeling and investigations for optimal electrode montage. Expert Rev Neurother 19:1025–1035.

17. Everitt BJ, Robbins TW (2016) Drug Addiction: Updating Actions to Habits to Compulsions Ten Years On. Annual Review of Psychology 67:23–50.

18. Fregni F, Pascual-Leone A (2007) Technology Insight: noninvasive brain stimulation in neurology[mdash]perspectives on the therapeutic potential of rTMS and tDCS. Nat Clin Pract Neuro 3:383–393.

19. Fregni F, El-Hagrassy MM, Pacheco-Barrios K, Carvalho S, Leite J, Simis M, Brunelin J, Nakamura-Palacios EM, Marangolo P, Venkatasubramanian G, San-Juan D, Caumo W, Bikson M, Brunoni AR, Group NCW (2020) Evidence-Based Guidelines and Secondary Meta-Analysis for the Use of Transcranial Direct Current Stimulation in Neurological and Psychiatric Disorders. International Journal of Neuropsychopharmacology 24:256–313.

20. Gandiga PC, Hummel FC, Cohen LG (2006) Transcranial DC stimulation (tDCS): A tool for double-blind sham-controlled clinical studies in brain stimulation. Clinical Neurophysiology 117:845–850.

21. Gao T, Du J, Tian S, Liu W (2022) A meta-analysis of the effects of non-invasive brain stimulation on obsessive-compulsive disorder. Psychiatry Research 312:114530.

22. Goodman WK, Price LH, Rasmussen SA, et al. (1989) The yale-brown obsessive compulsive scale: I. development, use, and reliability. Archives of General Psychiatry 46:1006–1011.

23. Gowda SM, Narayanaswamy JC, Hazari N, Bose A, Chhabra H, Balachander S, Bhaskarapillai B, Shivakumar V, Venkatasubramanian G, Reddy YCJ (2019) Efficacy of pre-supplementary motor area transcranial direct current stimulation for treatment resistant obsessive compulsive disorder: A randomized, double blinded, sham controlled trial. Brain Stimulation 12:922–929.

24. Grabner G, Janke AL, Budge MM, Smith D, Pruessner J, Collins DL (2006) Symmetric Atlasing and Model Based Segmentation: An Application to the Hippocampus in Older Adults. In, pp 58-66. Berlin, Heidelberg: Springer Berlin Heidelberg.

25. Graybiel AM, Rauch SL (2000) Toward a Neurobiology of Obsessive-Compulsive Disorder. Neuron 28:343–347.

26. Hampshire A, Sharp DJ (2015) Contrasting network and modular perspectives on inhibitory control. Trends in Cognitive Sciences 19:445–452.

27. Heinzel S, Kaufmann C, Grützmann R, Hummel R, Klawohn J, Riesel A, Bey K, Lennertz L, Wagner M, Kathmann N (2018) Neural correlates of working memory deficits and associations to response inhibition in obsessive compulsive disorder. NeuroImage: Clinical 17:426–434.

28. Ho K-A, Taylor JL, Chew T, Gálvez V, Alonzo A, Bai S, Dokos S, Loo CK (2016) The Effect of Transcranial Direct Current Stimulation (tDCS) Electrode Size and Current Intensity on Motor Cortical Excitability: Evidence From Single and Repeated Sessions. Brain Stimulation 9:1–7.

29. Huang Y, Datta A, Bikson M, Parra LC (2019) Realistic volumetric-approach to simulate transcranial electric stimulation—ROAST—a fully automated open-source pipeline. Journal of neural engineering 16:056006.

30. Hung Y, Gaillard SL, Yarmak P, Arsalidou M (2018) Dissociations of cognitive inhibition, response inhibition, and emotional interference: Voxelwise ALE meta-analyses of fMRI studies. Hum Brain Mapp 39:4065–4082.

31. Jafari E, Alizadehgoradel J, Pourmohseni Koluri F, Nikoozadehkordmirza E, Refahi M, Taherifard M, Nejati V, Hallajian A-H, Ghanavati E, Vicario CM, Nitsche MA, Salehinejad MA (2021) Intensified electrical stimulation targeting lateral and medial prefrontal cortices for the treatment of social anxiety disorder: A randomized, double-blind, parallel-group, dose-comparison study. Brain Stimulation 14:974–986.

32. Jog MV, Wang DJJ, Narr KL (2019) A review of transcranial direct current stimulation (tDCS) for the individualized treatment of depressive symptoms. Personalized Medicine in Psychiatry 17–18:17- 22.

33. Johansen-Berg H, Behrens TEJ, Robson MD, Drobnjak I, Rushworth MFS, Brady JM, Smith SM, Higham DJ, Matthews PM (2004) Changes in connectivity profiles define functionally distinct regions in human medial frontal cortex. Proc Natl Acad Sci U S A 101:13335–13340.

34. Leyfer OT, Ruberg JL, Woodruff-Borden J (2006) Examination of the utility of the Beck Anxiety Inventory and its factors as a screener for anxiety disorders. Journal of Anxiety Disorders 20:444–458.

35. Li H, Hu X, Gao Y, Cao L, Zhang L, Bu X, Lu L, Wang Y, Tang S, Li B, Yang Y, Biswal BB, Gong Q, Huang X (2020) Neural primacy of the dorsolateral prefrontal cortex in patients with obsessive-compulsive disorder. NeuroImage: Clinical 28:102432.

36. Maust D, Cristancho M, Gray L, Rushing S, Tjoa C, Thase ME (2012) Chapter 13 - Psychiatric rating scales. In: Handbook of Clinical Neurology (Aminoff MJ, Boller F, Swaab DF, eds), pp 227-237: Elsevier.

37. Meier SM, Mattheisen M, Mors O, Schendel DE, Mortensen PB, Plessen KJ (2016) Mortality Among Persons With Obsessive-Compulsive Disorder in Denmark. JAMA Psychiatry 73:268–274.

38. Menzies L, Achard S, Chamberlain SR, Fineberg N, Chen C-H, del Campo N, Sahakian BJ, Robbins TW, Bullmore E (2007) Neurocognitive endophenotypes of obsessive-compulsive disorder. Brain 130:3223-3236.

39. Minarik T, Berger B, Althaus L, Bader V, Biebl B, Brotzeller F, Fusban T, Hegemann J, Jesteadt L, Kalweit L, Leitner M, Linke F, Nabielska N, Reiter T, Schmitt D, Spraetz A, Sauseng P (2016) The Importance of Sample Size for Reproducibility of tDCS Effects. Frontiers in Human Neuroscience 10.

40. Monte-Silva K, Kuo M-F, Hessenthaler S, Fresnoza S, Liebetanz D, Paulus W, Nitsche MA (2013) Induction of Late LTP-Like Plasticity in the Human Motor Cortex by Repeated Non-Invasive Brain Stimulation. Brain Stimulation 6:424–432.

41. Nikolin S, Moffa A, Razza L, Martin D, Brunoni AR, Palm U, Padberg F, Bennabi D, Haffen E, Blumberger DM, Salehinejad MA, Loo CK (2023) Time-course of the tDCS antidepressant effect: An individual participant data meta-analysis. Progress in Neuro-Psychopharmacology and Biological Psychiatry 125:110752.

42. Nitsche M, Paulus W (2000) Excitability changes induced in the human motor cortex by weak transcranial direct current stimulation. The Journal of Physiology 527:633–639.

43. Norman LJ, Carlisi C, Lukito S, Hart H, Mataix-Cols D, Radua J, Rubia K (2016) Structural and functional brain abnormalities in attention-deficit/hyperactivity disorder and obsessive-compulsive disorder: a comparative meta-analysis. JAMA psychiatry 73:815–825.

44. Norman LJ, Taylor SF, Liu Y, Radua J, Chye Y, De Wit SJ, Huyser C, Karahanoglu FI, Luks T, Manoach D, Mathews C, Rubia K, Suo C, van den Heuvel OA, Yücel M, Fitzgerald K (2019) Error Processing and Inhibitory Control in Obsessive-Compulsive Disorder: A Meta-analysis Using Statistical Parametric Maps. Biological Psychiatry 85:713–725.

45. Overbeek T, Schruers K, Vermetten E, Griez E (2002) Comorbidity of obsessive-compulsive disorder and depression: Prevalence, symptom severity, and treatment effect. The Journal of Clinical Psychiatry 63:1106–1112.

46. Perera MPN, Mallawaarachchi S, Bailey NW, Murphy OW, Fitzgerald PB (2023) Obsessive-compulsive disorder (OCD) is associated with increased electroencephalographic (EEG) delta and theta oscillatory power but reduced delta connectivity. Journal of Psychiatric Research 163:310–317.

47. Pinto BS, Cavendish BA, da Silva PHR, Suen PJC, Marinho KAP, Valiengo LdCL, Vanderhasselt M-A, Brunoni AR, Razza LB (2022) The Effects of Transcranial Direct Current Stimulation in Obsessive– Compulsive Disorder Symptoms: A Meta-Analysis and Integrated Electric Fields Modeling Analysis. Biomedicines 11:80.

48. Polania R, Kuo M-F, Nitsche MA (2021) Physiology of Transcranial Direct and Alternating Current Stimulation. In: Transcranial Direct Current Stimulation in Neuropsychiatric Disorders: Clinical Principles and Management (Brunoni AR, Nitsche MA, Loo CK, eds), pp 29–47. Cham: Springer International Publishing.

49. Polanía R, Nitsche MA, Ruff CC (2018) Studying and modifying brain function with non-invasive brain stimulation. Nat Neurosci 21:174–187.

50. Rehn S, Eslick GD, Brakoulias V (2018) A Meta-Analysis of the Effectiveness of Different Cortical Targets Used in Repetitive Transcranial Magnetic Stimulation (rTMS) for the Treatment of Obsessive-Compulsive Disorder (OCD). Psychiatr Q 89:645–665.

51. Reuman L, Jacoby RJ, Blakey SM, Riemann BC, Leonard RC, Abramowitz JS (2017) Predictors of illness anxiety symptoms in patients with obsessive compulsive disorder. Psychiatry Research 256:417–422.

52. Robbins TW, Vaghi MM, Banca P (2019) Obsessive-Compulsive Disorder: Puzzles and Prospects. Neuron 102:27–47.

53. Romanelli RJ, Wu FM, Gamba R, Mojtabai R, Segal JB (2014) Behavioral therapy and serotonin reuptake inhibitor pharmacotherapy in the treatment of obsessive-compulsive disorder: a systematic review and meta-analysis of head-to-head randomized controlled trials. Depress Anxiety 31:641–652.

54. Rostami R, Kazemi R, Jabbari A, Madani AS, Rostami H, Taherpour MA, Molavi P, Jaafari N, Kuo M-F, Vicario CM, Nitsche MA, Salehinejad MA (2020) Efficacy and clinical predictors of response to rTMS treatment in pharmacoresistant obsessive-compulsive disorder (OCD): a retrospective study. BMC Psychiatry 20:372.

55. Salehinejad MA, Ghanavati E, Rashid MHA, Nitsche MA (2021a) Hot and cold executive functions in the brain: A prefrontal-cingular network. Brain and Neuroscience Advances 5:23982128211007769.

56. Salehinejad MA, Ghanavati E, Glinski B, Hallajian A-H, Azarkolah A (2022a) A systematic review of randomized controlled trials on efficacy and safety of transcranial direct current stimulation in major neurodevelopmental disorders: ADHD, autism, and dyslexia. Brain and Behavior 12:e2724.

57. Salehinejad MA, Wischnewski M, Ghanavati E, Mosayebi-Samani M, Kuo M-F, Nitsche MA (2021b) Cognitive functions and underlying parameters of human brain physiology are associated with chronotype. Nature Communications 12:4672.

58. Salehinejad MA, Ghanavati E, Reinders J, Hengstler JG, Kuo M-F, Nitsche MA (2022b) Sleep-dependent upscaled excitability, saturated neuroplasticity, and modulated cognition in the human brain. eLife 11:e69308.

59. Salehinejad MA, Nejati V, Mosayebi-Samani M, Mohammadi A, Wischnewski M, Kuo M-F, Avenanti A, Vicario CM, Nitsche MA (2020) Transcranial Direct Current Stimulation in ADHD: A Systematic Review of Efficacy, Safety, and Protocol-induced Electrical Field Modeling Results. Neurosci Bull 36:1191–1212.

60. Schroeder PA, Schwippel T, Wolz I, Svaldi J (2020) Meta-analysis of the effects of transcranial direct current stimulation on inhibitory control. Brain Stimulation: Basic, Translational, and Clinical Research in Neuromodulation 13:1159–1167.

61. Sharp DJ, Bonnelle V, De Boissezon X, Beckmann CF, James SG, Patel MC, Mehta MA (2010) Distinct frontal systems for response inhibition, attentional capture, and error processing. Proceedings of the National Academy of Sciences 107:6106–6111.

62. Shin NY, Lee TY, Kim E, Kwon JS (2013) Cognitive functioning in obsessive-compulsive disorder: a meta-analysis. Psychological Medicine 44:1121–1130.

63. Silva RdMFd, Brunoni AR, Goerigk S, Batistuzzo MC, Costa DLdC, Diniz JB, Padberg F, D’Urso G, Miguel EC, Shavitt RG (2021) Efficacy and safety of transcranial direct current stimulation as an add-on treatment for obsessive-compulsive disorder: a randomized, sham-controlled trial. Neuropsychopharmacology 46:1028–1034.

64. Skevington SM, Lotfy M, O’Connell KA (2004) The World Health Organization’s WHOQOL-BREF quality of life assessment: Psychometric properties and results of the international field trial. A Report from the WHOQOL Group. Qual Life Res 13:299–310.

65. Stagg CJ, Nitsche MA (2011) Physiological Basis of Transcranial Direct Current Stimulation. The Neuroscientist 17:37–53.

66. Steer RA, Beck AT (1997) Beck Anxiety Inventory. In: Evaluating stress: A book of resources., pp 23-40. Lanham, MD, US: Scarecrow Education.

67. Stein DJ, Costa DLC, Lochner C, Miguel EC, Reddy YCJ, Shavitt RG, van den Heuvel OA, Simpson HB (2019) Obsessive–compulsive disorder. Nature Reviews Disease Primers 5:52.

68. Stella J. de Wit, M.D. ,, Froukje E. de Vries, M.D. ,, Ysbrand D. van der Werf, Ph.D. ,, Danielle C. Cath, M.D., Ph.D. ,, Dirk J. Heslenfeld, Ph.D. ,, Eveline M. Veltman, M.Sc. ,, Anton J.L.M. van Balkom, M.D., Ph.D. ,, Dick J. Veltman, M.D., Ph.D. , and, Odile A. van den Heuvel, M.D., Ph.D. (2012) Presupplementary Motor Area Hyperactivity During Response Inhibition: A Candidate Endophenotype of Obsessive-Compulsive Disorder. American Journal of Psychiatry 169:1100–1108.

69. van den Heuvel OA, Veltman DJ, Groenewegen HJ, Witter MP, Merkelbach J, Cath DC, van Balkom AJLM, van Oppen P, van Dyck R (2005) Disorder-Specific Neuroanatomical Correlates of Attentional Bias in Obsessive-compulsive Disorder, Panic Disorder, and Hypochondriasis. Archives of General Psychiatry 62:922–933.

70. van den Heuvel OA, van Wingen G, Soriano-Mas C, Alonso P, Chamberlain SR, Nakamae T, Denys D, Goudriaan AE, Veltman DJ (2016) Brain circuitry of compulsivity. Eur Neuropsychopharmacol 26:810–827.

71. Velikova S, Locatelli M, Insacco C, Smeraldi E, Comi G, Leocani L (2010) Dysfunctional brain circuitry in obsessive–compulsive disorder: Source and coherence analysis of EEG rhythms. Neuroimage 49:977–983.

72. Velloso P, Piccinato C, Ferrão Y, Aliende Perin E, Cesar R, Fontenelle L, Hounie AG, do Rosário MC (2016) The suicidality continuum in a large sample of obsessive–compulsive disorder (OCD) patients. Eur Psychiatry 38:1–7.

73. Wayne K. Goodman, M.D. ,, Eric A. Storch, Ph.D. ,, Sameer A. Sheth, M.D., Ph.D. (2021) Harmonizing the Neurobiology and Treatment of Obsessive-Compulsive Disorder. American Journal of Psychiatry 178:17–29.

74. Wong M, Woody EZ, Schmidt LA, Ameringen MV, Soreni N, Szechtman H (2015) Frontal EEG alpha activity and obsessive-compulsive behaviors in non-clinical young adults: a pilot study. Front Psychol 6:1480.

75. Woods AJ, Antal A, Bikson M, Boggio PS, Brunoni AR, Celnik P, Cohen LG, Fregni F, Herrmann CS, Kappenman ES, Knotkova H, Liebetanz D, Miniussi C, Miranda PC, Paulus W, Priori A, Reato D, Stagg C, Wenderoth N, Nitsche MA (2016) A technical guide to tDCS, and related non-invasive brain stimulation tools. Clinical Neurophysiology 127:1031–1048.

